# Association of Complement C3 Inhibitor Pegcetacoplan with Photoreceptor Degeneration Beyond Areas of Geographic Atrophy

**DOI:** 10.1101/2022.06.20.22276634

**Authors:** Maximilian Pfau, Steffen Schmitz-Valckenberg, Ramiro Ribeiro, Reza Safaei, Alex McKeown, Monika Fleckenstein, Frank G. Holz

## Abstract

Preservation of photoreceptors beyond areas of retinal pigment epithelium atrophy is a critical treatment goal in eyes with geographic atrophy (GA) to prevent vision loss. Thus, we assessed the association of treatment with the complement C3 inhibitor pegcetacoplan with optical coherence tomography (OCT)-based photoreceptor laminae thicknesses in this post hoc analysis of the FILLY trial (NCT02503332).

Retinal layers in OCT were segmented using a deep-learning-based pipeline and extracted along evenly spaced contour-lines surrounding areas of GA. The primary outcome measure was change from baseline in (standardized) outer nuclear layer (ONL) thickness at the 5.16°-contour-line at month 12.

Participants treated with pegcetacoplan monthly had a thicker ONL along the 5.16° contour-line compared to the pooled sham arm (mean difference [95% CI] +0.29 z-score units [0.16, 0.42],P<.001). The same was evident for eyes treated with pegcetacoplan every other month (+0.26 z-score units [0.13, 0.4],P<.001). Additionally, eyes treated with pegcetacoplan exhibited a thicker photoreceptor inner segment layer along the 5.16°-contour-line at month 12.

These findings suggest that pegcetacoplan could slow GA progression and lead to a lesser thinning of photoreceptor layers beyond the GA boundary. Future trials in earlier disease stages, i.e., intermediate AMD, aiming to slow photoreceptor degeneration warrant consideration.

## INTRODUCTION

Geographic atrophy (GA), the atrophic late-stage manifestation of age-related macular degeneration, is a leading cause of legal blindness in industrialized countries.^1–3^ To date, no treatment options – beyond low vision aids – are available. Multiple complement inhibitors are currently evaluated in clinical trials. These include pegcetacoplan (APL-2), which slowed GA progression in recent Phase 2 and 3 trials.^4–6^

The defining lesions of GA are foci of retinal pigment epithelium (RPE), photoreceptor, and choriocapillaris atrophy that progress and fuse over time.^7^ The area of these lesions is associated with a corresponding visual function loss,^8,9^ impairment of activities of daily living,^10^ and reduced quality of life.^11,12^ Thus, the total area of RPE-atrophy has served as the primary outcome measure in multiple trials.

However, eyes with GA are also affected by photoreceptor degeneration beyond areas of RPE-atrophy. This photoreceptor degeneration is observable as localized thinning of the outer nuclear layer (ONL) in the so-called junctional zone (500 µm band surrounding GA).^13,14^ In addition, photoreceptor degeneration may be more widespread, especially in association with subretinal drusenoid deposits (SDD, or reticular pseudodrusen).^15–17^ This more diffuse form of outer retinal degeneration can be quantified as an outer nuclear layer (ONL), photoreceptor inner segment (IS), and outer segment (OS) thinning in eyes with intermediate AMD,^17–19^ and GA.^14,20–22^ Importantly, thinning at the level of the ONL is an established surrogate of impaired light sensitivity.^17–19,23^ A recent analysis showed that treatment with pegcetacoplan is associated with less thinning between the inner boundary of the ellipsoid zone and the outer boundary of band three. However, data on the ONL and differential data for the IS and OS layers are lacking to date.^24^

Thus, we aimed to assess the association of pegcetacoplan treatment with the change individual photoreceptor laminae thicknesses in patients with GA secondary to AMD. The presented analysis using data from the FILLY phase 2 trial (ClinicalTrials.gov identifier NCT02503332) fully accounts for the normal spatial variation in photoreceptor laminae thicknesses aiming to evaluate photoreceptor degeneration independent of the changes related to mere differences in RPE-atrophy progression rates.

## RESULTS

### Cohort

Forty-nine of the 246 clinical trial participants were excluded since they underwent spectral-domain optical coherence tomography (SD-OCT) imaging using a Cirrus device (Supplementary Figure S1).

A total of 197 participants were imaged with a Heidelberg Spectralis OCT device, of whom 192 had follow-up imaging data available and met the prespecified modified intention-to-treat (mITT) criteria for analysis (i.e., 97.5 % of the participants imaged with a Heidelberg Spectralis OCT device, and 78 % of all randomized participants). The participants (female: 122 [63.5%]; male: 70 [36.5%]) had a mean age of 79.4 years (SD: 7.53; range: 60.0 to 97.0) at baseline with a mean (autofluorescence based) sqrt-transformed area of fundus autofluorescence (FAF)-based RPE-atrophy of 2.80 mm (SD: 0.718; range: 1.59, 4.16). The three study arms had no distinct differences (Table 1).

**Table 1.**
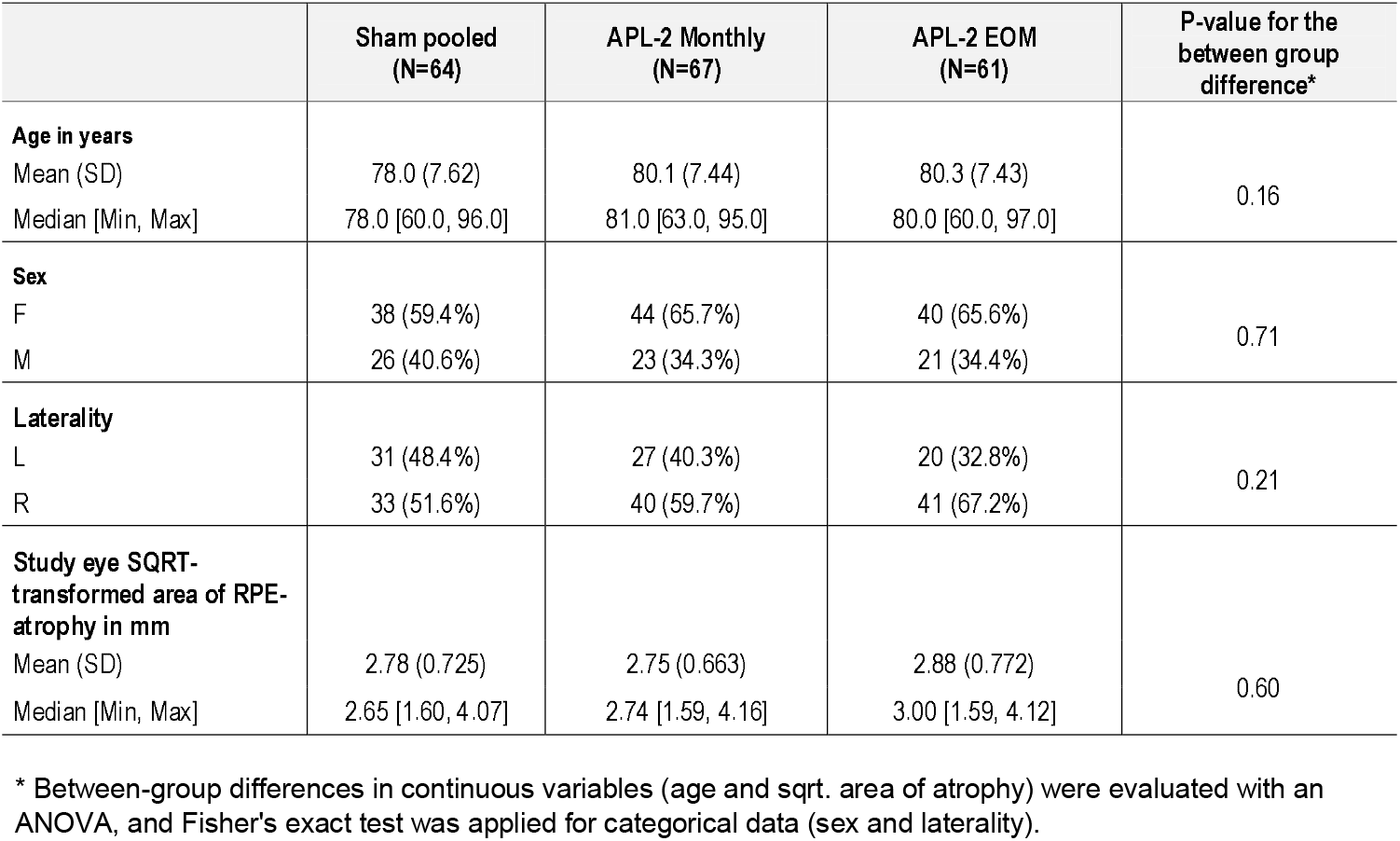
Cohort characteristics for the included participants (modified intention-to-treat [mITT] with Spectralis imaging) ***Abbreviations:*** *retinal pigment epithelium (RPE), square-root (SQRT)*

Participants enrolled at clinical sites using the Cirrus device, which could not be included in this analysis, were well-aligned with the included participants from sites using the Spectralis device (Supplementary Table S1)

### Segmentation Accuracy

To assess the accuracy of the segmentation pipeline, we compared our SD-OCT-derived measurement of the area of RPE-atrophy (cf., Figure 1) with the previously reported FAF-based measurement of RPE-atrophy. For the subset of study eyes with measurable RPE-atrophy (i.e., RPE-atrophy within the SD-OCT image frame), there was no relevant difference (systematic bias) between the original FAF-based measurements and the SD-OCT derived data (bias estimate [95% CI] of 0.02 mm [0.00; 0.05]. P=0.020, Supplementary Figure S2).

**Figure 1.**
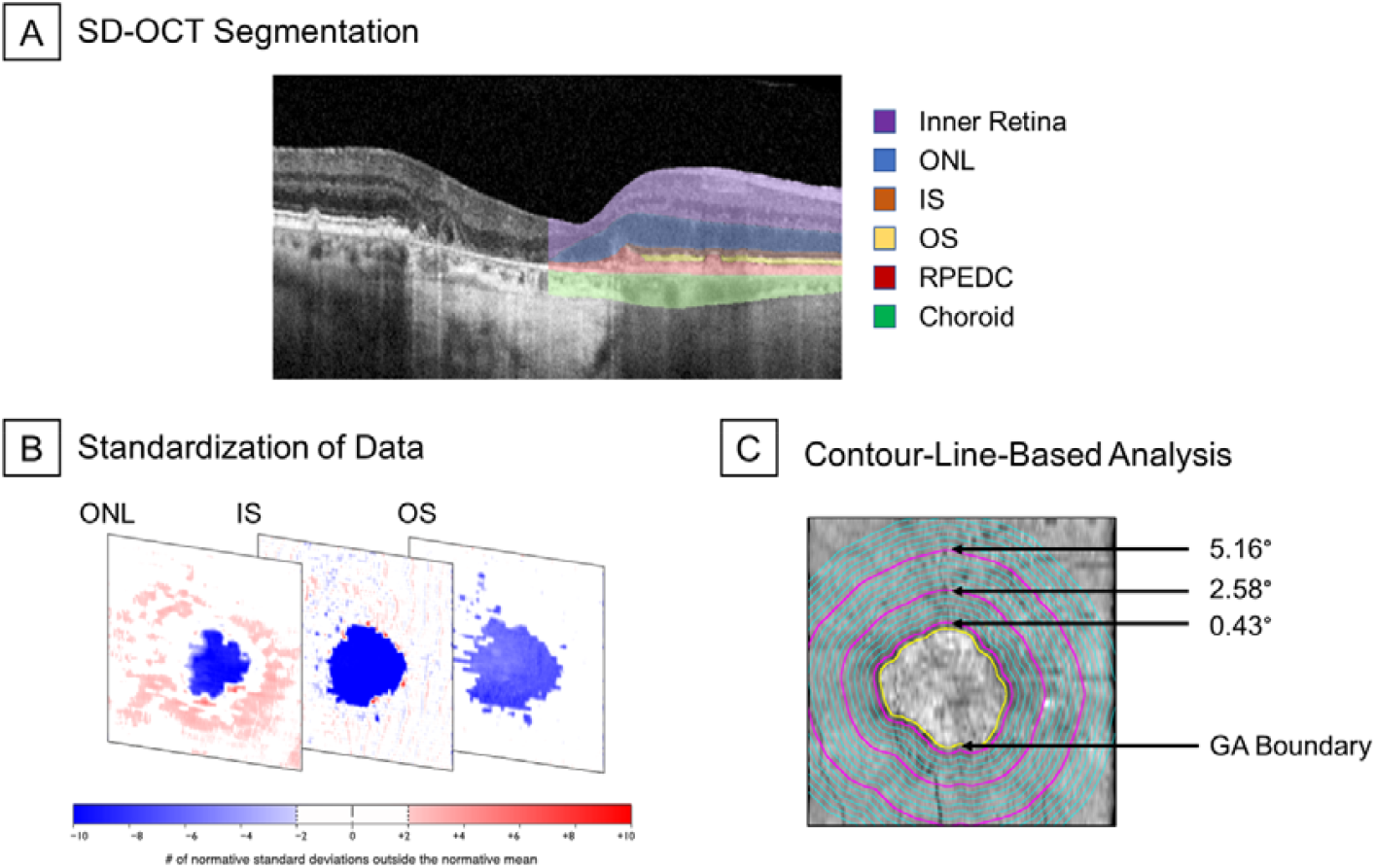
Image segmentation approach. As shown in the first panel, all spectral-domain optical coherence tomography (SD-OCT) data were segmented using a convolutional neural network (CNN, Deeplabv3 model with a ResNet-50 backbone). Subsequently (second panel), these data were standardized (z-scores) to account for normal thickness variation due to age and retinal topography. Last (third panel), the retinal layers’ thicknesses were extracted along evenly spaced contour-lines surrounding the retinal pigment epithelium (RPE) atrophy area. The yellow line denotes the boundary of RPE atrophy. The three purple lines represent the 0.43°, 2.58°, and 5.16° contour-lines. ***Abbreviations:*** *spectral-domain optical coherence tomography (SD-OCT), convolutional neural network (CNN), retinal pigment epithelium (RPE), outer nuclear layer (ONL), photoreceptor inner segments (IS), photoreceptor outer segments (OS), retinal pigment epithelium-drusen complex (RPEDC)*

### SD-OCT-based Trial Outcome

The primary outcome measure for the SD-OCT-based assessment mirrored the previous FAF-based report. Specifically, for the subset of study eyes with GA within the image frame (n = 135), the square-root transformed atrophy progression was slower by 25% in eyes treated with pegcetacoplan monthly (sham - pegcetacoplan monthly estimate: +0.076 mm/y, SE: 0.038, P=0.0473), and in eyes treated with pegcetacoplan every other month (EOM) 22.2% (sham pooled - pegcetacoplan EOM estimate: +0.068 mm/y, SE: 0.04, P=.09).

### Photoreceptor Layer Thickness Outside of GA

At baseline, all three arms (sham pooled, pegcetacoplan monthly, and pegcetacoplan EOM) exhibited similar degrees of photoreceptor thinning. Specifically, the ONL was thinned in the immediate junctional zone, with a steep gradient between 0° to 2° [approx. 0 to 582 µm] from the boundary of RPE-atrophy and a less steep gradient at more eccentric locations (Figure 2). Similarly, marked IS and OS thinning was evident outside of RPE-atrophy across all groups at baseline (Supplementary Figure S3).

**Figure 2.**
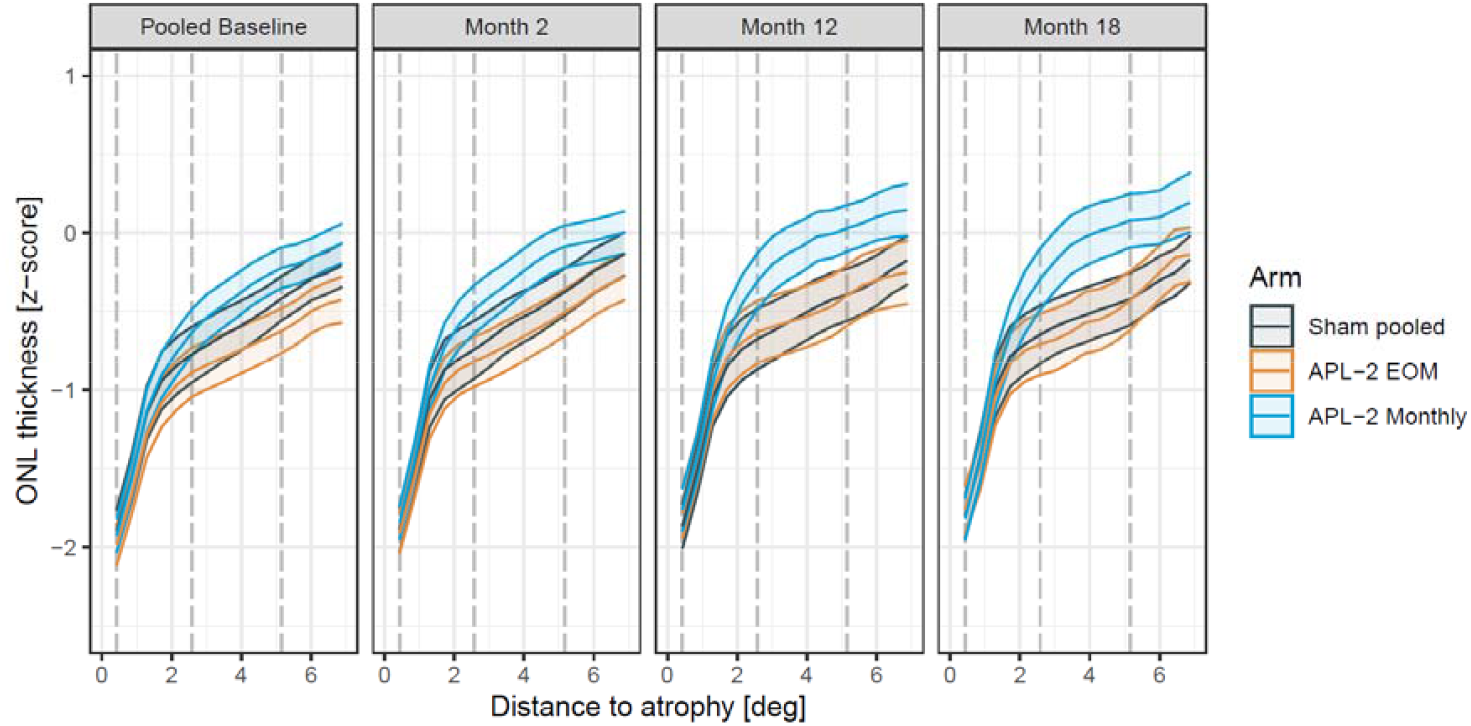
Change in thickness at the level of the outer nuclear layer (ONL) over time. The plots show the average thickness at the level of the outer nuclear layer (ONL, y-axis) and the standard error of the mean (SEM, ribbons) with the distance to the boundary of retinal pigment epithelium (RPE) atrophy (x-axis). The panels indicate the visit, and the color indicates the treatment arm. The vertical dashed lines denote the 0.43°, 2.58°, and 5.16° contour-lines that were considered for the linear mixed model analyses. Please note that the baseline junctional-zone ONL thickness differed slightly among the groups. The actual contrasts (i.e., change-over-time from baseline) are shown in Figure 3. The data were derived from the modified intention-to-treat (mITT) analysis (N_participants_= 192). ***Abbreviations:*** *outer nuclear layer (ONL), photoreceptor inner segments (IS), photoreceptor outer segments (OS), modified intention-to-treat (mITT)*

### Change in Photoreceptor Layer Thickness Over Time

At month 12, eyes treated with pegcetacoplan exhibited a lesser degree of progressive thinning at the level of the ONL along their new junctional zone compared to eyes in the sham arm. Specifically, the ONL along the 5.16° contour-line was markedly thicker compared to sham in the pegcetacoplan monthly group (contrast estimate [95% CI] for pegcetacoplan monthly - sham: +0.29 z-score units [0.16, 0.42], P<.001) and in the pegcetacoplan EOM group (pegcetacoplan EOM - sham: +0.26 z-score units [0.13, 0.4], P<.001).

Likewise, the retina at the level of the IS layer was also thicker in both treatment arms compared to sham at month 12 along the 5.16° contour-line (pegcetacoplan monthly - sham: +0.42 z-score units [0.21, 0.62], P<.001; pegcetacoplan EOM sham: +0.34 z-score units [0.12, 0.55], P<.001). The same was observed for the 2.58° contour-line (Table 2, Supplementary Figure S4).

**Table 2.** Study eye differences in thickness at the level of photoreceptor layers (in z-score units) at three contour-lines at month 12 (modified intention-to-treat [mITT] analysis)

| Layer | Contrast | 0.43° |  |  | 2.58° |  |  | 5.16° |  |  |
| --- | --- | --- | --- | --- | --- | --- | --- | --- | --- | --- |
|  |  | Estimate | 95% CI | P-value* | Estimate | 95% CI | P-value* | Estimate | 95% CI | P-value* |
| ONL | (APL-2 Monthly) - Sham pooled | 0.1 | [-0.03, 0.24] | .16 | 0.26 | [0.14, 0.38] | <.001 | 0.29 | [0.16, 0.42] | <.001 |
| ONL | (APL-2 EOM) - Sham pooled | 0.12 | [-0.01, 0.26] | .09 | 0.16 | [0.04, 0.29] | .006 | 0.26 | [0.13, 0.4] | <.001 |
| ONL | (APL-2 Monthly) - (APL-2 EOM) | -0.02 | [-0.15, 0.12] | .95 | 0.09 | [-0.03, 0.22] | .17 | 0.02 | [-0.11, 0.16] | .92 |
| IS | (APL-2 Monthly) - Sham pooled | 0.68 | [0.09, 1.28] | .02 | 0.48 | [0.21, 0.75] | <.001 | 0.42 | [0.21, 0.62] | <.001 |
| IS | (APL-2 EOM) - Sham pooled | 1.09 | [0.48, 1.71] | <.001 | 0.4 | [0.12, 0.68] | .002 | 0.34 | [0.12, 0.55] | <.001 |
| IS | (APL-2 Monthly) - (APL-2 EOM) | -0.41 | [-1.02, 0.2] | .26 | 0.08 | [-0.2, 0.36] | .78 | 0.08 | [-0.13, 0.29] | .64 |
| OS | (APL-2 Monthly) - Sham pooled | -0.09 | [-0.38, 0.19] | .72 | 0.18 | [-0.12, 0.48] | .33 | 0.29 | [-0.01, 0.58] | .06 |
| OS | (APL-2 EOM) - Sham pooled | 0.05 | [-0.24, 0.35] | .9 | 0.52 | [0.21, 0.83] | <.001 | 0.3 | [-0.01, 0.6] | .06 |
| OS | (APL-2 Monthly) - (APL-2 EOM) | -0.15 | [-0.44, 0.14] | .45 | -0.34 | [-0.64, -0.03] | .03 | -0.01 | [-0.31, 0.3] | 1 |

\* P-values were obtained using Kenward-Roger approximation to estimate the denominator degrees of freedom. P-values were adjusted within each model (i.e., combination of layer and contour-line) using the Tukey method for comparing a family of 3 estimates.

At the level of the OS layer, the results were overall indistinct (Table 2). The OS layer differed at month 12 only along the 2.58° contour-line markedly in thickness in the pegcetacoplan EOM group compared to sham (pegcetacoplan EOM - sham: +0.52 z-score units [0.21, 0.83], P<.001).

Analysis of the change over time (Figure 3), revealed for the ONL and IS less thinning at the level of the photoreceptors with increasing duration of treatment (i.e., estimate at month 12 and 6 > month 2) and with more frequent dosing (estimates for pegcetacoplan monthly > pegcetacoplan EOM). Notably, these differences were evident through month 18. The PP analysis (Supplementary Table S2, Supplementary Figure S5) and the PP analysis excluding all visits from eyes with exudation at any time point (Supplementary Table S3, Supplementary Figure S6) confirmed these results with even larger coefficients.

**Figure 3.**
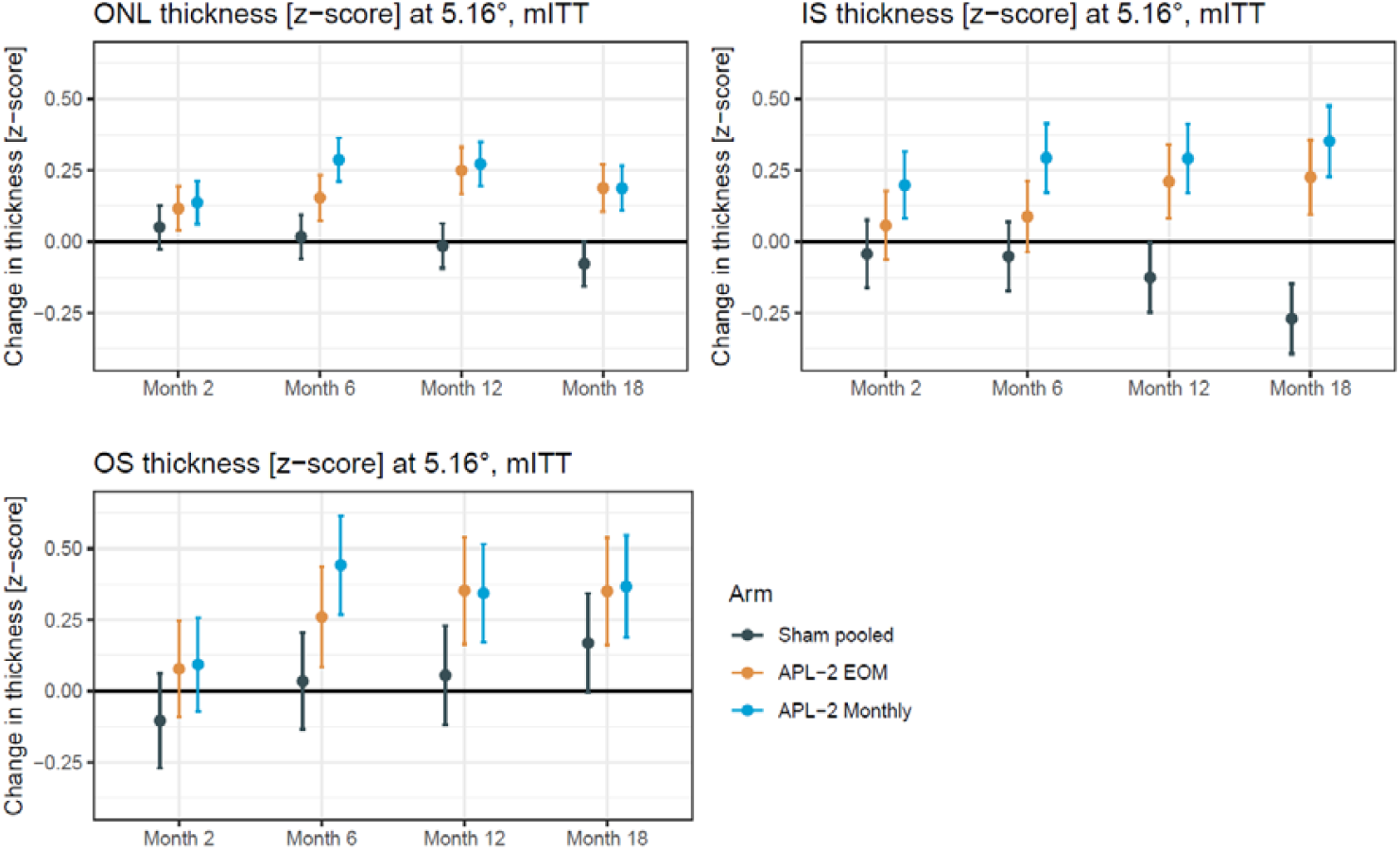
Change in thickness at the level of the photoreceptor layers along the 5.16° contour-line over time, modified intention-to-treat (mITT) analysis. The plots show the least-squares (adjusted) means from the linear mixed model analysis for the change in thickness at the level of the outer nuclear layer (ONL), photoreceptor inner segments (IS), and photoreceptor outer segments (OS) along the 5.16° contour-line as a function of the visit (x-axis) and treatment arm (colors). The vertical lines denote the 95% confidence intervals. Eyes treated with pegcetacoplan (APL-2) monthly tended to show a lesser degree of photoreceptor laminae thinning over time. Participants were treated between baseline and month 12. The data were derived from the modified intention-to-treat (mITT) analysis (N_participants_= 192). ***Abbreviations:*** *outer nuclear layer (ONL), photoreceptor inner segments (IS), photoreceptor outer segments (OS), modified intention-to-treat (mITT)*

### Association of Pegcetacoplan with Fellow Eye Photoreceptor Thickness

Fellow eye data (eyes without macular neovascularization and a baseline RPE-atrophy area ≥ 2.5 mm^2^) were available for 112 participants (eligible fellow eyes per arm: sham N=41, pegcetacoplan monthly N=33, pegcetacoplan EOM N=38). Applying the same contour-line-based analysis in these eyes yielded similar results concerning baseline thinning of the ONL, IS, and OS in the junctional zone. However, there were no longitudinal changes in the fellow eyes of all three arms (Supplementary Table S4, Supplementary Figure S7).

## DISCUSSION

This study analyzed the impact of the C3 inhibitor pegcetacoplan on photoreceptor degeneration outside of RPE-atrophy using data from the randomized FILLY phase-2 trial. Specifically, we demonstrated that eyes treated with pegcetacoplan exhibit a lesser degree of thinning at the level of photoreceptors outside of RPE-atrophy at the end of the study compared to eyes in the sham arm.

The area of RPE-atrophy has been established as the primary structural endpoint to quantify disease progression in eyes with GA.^2^ For this purpose, RPE-atrophy is well suited since it represents the boundary of deep scotomata in eyes with GA as shown by fundus-controlled perimetry studies,^8,26^ is prognostic for reading ability,^27–29^ as for quality of life.^11,27^ While early studies typically used fundus photography for the quantification of the area of RPE-atrophy,^30^ FAF-based quantification using semiautomated software such as the RegionFinder software (Heidelberg Engineering, Germany) is currently the gold standard used in large-scale clinical trials^31–33^ More recently, GA quantification with other semi and fully automated approaches for FAF or OCT data has been proposed.^34–37^ However, the area of RPE-atrophy depicts only partially the severity of AMD in a patient with GA.

In the setting of intermediate AMD and GA, it is now established that photoreceptors – especially in association with SDD – show degenerative changes over time in a macula-wide manner.^15,17,20,21,38^ Importantly, histopathologic data are congruent with photoreceptor degeneration distant to the boundary of RPE-atrophy,^14^ and suggest that rod photoreceptor degeneration precedes cone degeneration in AMD.^39,40^ Therefore, treatment of the non-exudative component of AMD should ideally address not only RPE-atrophy but also photoreceptor degeneration in the broader sense. Thus, we have analyzed the association of pegcetacoplan with photoreceptor degeneration outside of RPE-atrophy.

Interestingly, the analyses for all three photoreceptor laminae demonstrated that pegcetacoplan was associated with less thinning at the level of photoreceptors outside areas of RPE-atrophy. Considering Bradford Hill’s criteria for causation,^41^ the here observed associations may be causal. First and foremost, the association strength is substantial, follows a plausible temporal sequence (estimate increases over time), and shows a biological gradient (dose-response relationship with a more substantial impact for eyes treated monthly instead of EOM). Moreover, the association is specific, as evidenced by the absence of an association in the fellow eyes. Since photoreceptor thinning correlates closely to impaired light sensitivity,^17,19,42–44^ it is possible that the observed changes are functionally beneficial.

Overall, the results shown here align with the previous analysis by Sophie Riedl and coworkers.^24^ Through differential analysis of all three photoreceptor laminae, our data suggests that their observed effect (based on a combined IS+OS layer definition) is predominantly attributable to changes at the IS level. Importantly, our analysis fully accounted for the spatial variation in photoreceptor laminae thickness through standardization. Thus, our results attest that the preservation of photoreceptor laminae thickness in the junctional zone can not be attributed to an overall less eccentric junctional zone in treated eyes at month 12 due to the slower RPE-atrophy progression. Instead, the observation presents a genuine lesser degree of photoreceptor thinning independent of the underlying RPE-atrophy progression.

As a related concept, Wu and coworkers have proposed ‘nascent GA’, a combination of OCT imaging signs preceding RPE-atrophy, as an endpoint applicable to evaluate retinal degeneration in eyes with intermediate AMD,^45^ and demonstrated its prognostic value.^46^ This concept was more recently incorporated by the Classification of Atrophy Meeting (CAM) consortium as a part of incomplete RPE and outer retinal atrophy (iRORA).^47,48^ Besides incident iRORA (or nGA) as an endpoint, the rate of iRORA to RPE-atrophy transition distant to the junctional zone has been proposed as an endpoint.^49^ In another post-hoc study of the FILLY trial, a reduced iRORA to RPE-atrophy transition rate in eyes treated with pegcetacoplan was reported.^50^ Further studies are warranted to examine whether the observed lesser thinning at the level of the ONL along contour-lines reflects the sum of decreased focal thinning primarily,^50^ or reduced thinning in a ‘macula-wide’ manner.

### Limitations

This analysis was a post-hoc study. Accordingly, the results must be considered hypothesis-generating rather than confirming.

The approach of using ‘traveling’ contour-lines (after standardization of the thickness data) entails assumptions. This includes the absence of spatial patterns of (marked) photoreceptor degeneration unrelated to the junctional zone. The relative stability (or trend toward slight thinning) of the junctional zone photoreceptor thicknesses over time in the here presented fellow eye data and previous natural-history data implies that these assumptions are met.^20^

Notably, functional evidence will be needed to demonstrate genuine photoreceptor protection with absolute certainty (instead of the unlikely case of subtle thickening due to other causes).^51^ There is no evidence that the here observed findings are a result of ‘pre- or subclinical exudation’, given that the analysis excluding all visits from eyes that developed exudation at any time point showed consistent results (Supplementary Table S3, Supplementary Figure S6).

It is unclear why IS thickness showed the most distinct change in treated eyes. Histopathologic data would suggest that both IS and OS thickness are suitable biomarkers for photoreceptor integrity. In contrast, ONL thickness is partially confounded by HFL.^14^ Ultrahigh-resolution OCT technology would most likely provide more precise estimates of change-over-time in IS and OS thickness given the greater axial resolution.^52^ Last, analysis of photoreceptor degeneration outside of RPE-atrophy in the larger phase 3 trials investigating the same drug is warranted (i.e., DERBY, OAKS) and across investigational drugs (including among other avacincaptad pegol,^53^ IONIS-FB-LRx,^54^ FHTR2163^55^).

## CONCLUSION

In summary, this post-hoc analysis of the FILLY trial showed that treatment with pegcetacoplan was associated with less outer retinal thinning over time in a dose-dependent manner. Specifically, eyes treated with monthly pegcetacoplan (compared to controls) exhibited at month 12 relatively more intact tissue at the level of the ONL, IS, and OS laminae outside of the RPE-atrophy junctional zone. These results may support a therapeutic effect of pegcetacoplan on photoreceptors in eyes with GA.

## METHODS

### Clinical Trial Data

This post-hoc analysis was conducted between August 2021 and February 2022 and based on data acquired previously in the FILLY trial and is reported with adherence to the CONSORT standard. The protocol has been described previously (study start date: September 24, 2015, primary completion date: July 14, 2017).^5^ Ethics approval was obtained from all necessary boards, written informed consent was obtained from all participants, and participants did not receive a stipend.^5^ Sex was self-reported.

FILLY was a multicenter, randomized, single-masked, sham-controlled study to assess the safety of pegcetacoplan in patients with GA secondary to AMD. Study participants had to have a total GA area ≥ 2.5 mm^2^ (with one focus of GA ≥ 1.25 mm^2^ in eyes with multifocal GA) and ≤ 17.5 mm^2^ as assessed by fundus autofluorescence imaging. A total of 246 participants were included and randomized in a 2:2:1:1 manner to pegcetacoplan monthly, pegcetacoplan every other month (EOM), sham monthly, or sham EOM.^5^

Study participants in the monthly group received pegcetacoplan (or sham) injections and study procedures monthly until month 12. Participants in the EOM groups received pegcetacoplan (or sham) injections every two months until month 12. In addition, all participants were imaged at a month 15 and month 18 follow-up visit (3 and 6 months after the last injection, respectively).^5^

### Imaging Data

Besides fundus autofluorescence (FAF) imaging, participants underwent (among other imaging modalities) SD-OCT imaging, either with a Spectralis device (20°x20°, 49 B-scans, N=197 participants, Heidelberg Engineering, Heidelberg, Germany) or with a Cirrus device (N=49 participants, Carl Zeiss Meditec, Jena, Germany). The first group (i.e., participants imaged with a Spectralis device) was included in this analysis (Supplementary Figure S1). Data from five participants were excluded due to lack of follow-up or deviation from the SD-OCT protocol. Data acquired with the Cirrus device were excluded due to the lower axial resolution.

### Prespecified Study Population and Pooling of the Sham Arms

In analogy to the prespecified analysis of the primary endpoint,^5^ the modified intention-to-treat (mITT) population included all participants with at least one injection and at least one follow-up examination at month 2 (or later) at which efficacy data were collected. The per-protocol (PP) population included all participants from the mITT population that did receive no less than 75% of their expected injections before month 12 (i.e., no less than nine [monthly group] or four injections [EOM group]) and the participants that did not receive the incorrect medication throughout the study. We also report the results for the PP population, excluding all visits from eyes that developed exudation at any time point. Both sham arms were pooled for the analyses.^5^

### Image Data Segmentation and Standardization

A previously validated convolutional neural network (CNN)^20^ was applied to segment the retinal layers (cf., eMethods for layer definitions and segmentation methods). For the primary analyses, we compared photoreceptor laminae distances at fixed distances to the boundary of GA across time. Since these contour-lines ‘traveled’ with GA progression, all OCT thickness data were standardized to account for the normal topographic difference in photoreceptor laminae thickness and the effect of age. Specifically, we subtracted for each A-scan in the en-face maps the age-adjusted normal value of a given layer thickness and divided that value by the normal location-specific standard deviation (i.e., transformation to z-score units). This process has been described previously in detail.^20^

### Feature Extraction

The mean thickness values for the three photoreceptor laminae (ONL, IS, OS) were extracted along evenly spaced contour-lines around the lesions of GA (width: 0.43° [126 µm in an emmetropic eye]).^20^ The statistical analyses were performed for three representative contour-lines (distances to the GA boundary: 0.43°, 2.58°, and 5.16° [126 µm, 751 µm, and 1502 µm in an emmetropic eye]). These evenly spaced contour-lines were derived through dilation of the initial segmentation of GA. Outer contour-lines could be discontinuous due to the image-frame. Notably, the contour-lines were defined on the current (i.e., same visit date) position of the GA boundary for all visits. This means that the counter-lines ‘traveled’ as the lesions expanded. The generated data are thus independent of the underlying rate of RPE-atrophy progression (i.e., not a mere representation of the slowed rate of GA progression with treatment). In the absence of treatment effects, there should be a slight thinning of photoreceptor laminae over time due to a subtle, ‘macula-wide’ component of photoreceptor degeneration.^20^

### Statistical analyses

The analyses were performed in the software environment R (version 4.1.0),^56^ using the add on libraries dplyr (version 1.0.7),^57^ ggplot2 (version 3.3.5),^58^ lme4 (1.1-27.1),^59^ and emmeans (version 1.6.2-1).^60^

The primary outcome measure was the between-group difference in change from baseline of the ONL thickness along the 5.16° contour-line at month 12. This distance was elected as the primary outcome measure to be genuinely independent of the junctional-zone component of GA. We also examined two additional contour-lines (0.43°, 2.58°, besides 5.16°), and fitted for each photoreceptor layer (ONL, IS, OS), a linear mixed model (LMM) based on the observed data (i.e., no imputation). The dependent variable was the change in layer thickness (z-score) from baseline, and the independent variables (fixed effects) were the treatment arm (pegcetacoplan monthly, pegcetacoplan EOM, and pooled sham), baseline layer thickness, visit (month 2, 6, 12, 18), and the treatment arms by visit interaction. The model included the patient ID as a random effect. The Kenward-Roger approximation was used to estimate the denominator degrees of freedom. P-values were adjusted for the three pairwise contrasts between the three groups (Tukey’s single-step multiple comparison procedure). In this post-hoc analysis, estimates from a total of 27 LMMs are presented (3 [contour-lines] x 3 [layers: ONL, IS, OS] x 3 [analyses: mITT, PP, fellow eyes]).

As an additional quality-control of the OCT segmentation, we performed an OCT-based analysis of the effect of pegcetacoplan on RPE-atrophy progression rates in analogy to the autofluorescence-based analysis in the original publication.^5^ The change in square-root-transformed RPE-atrophy area from baseline was the dependent variable. The treatment arm (pegcetacoplan monthly, pegcetacoplan EOM, and pooled sham), baseline square-root-transformed RPE-atrophy area, visit (month 2, 6, 12, 18), and the treatment arms by visit interaction and the sqrt-baseline-area by visit interaction were included as the fixed effects. The model included the patient ID as a random effect. In addition, a Bland-Altman analysis was performed to compare the OCT and FAF data.^61^

## Data Availability

Data Sharing:
Original data will be shared from the corresponding authors upon reasonable request.

## ADDITIONAL INFORMATION

### Author Contributions

All authors are responsible for conception and design of the study. MP, and SSV are responsible for the analysis of the data. All authors contributed to interpretation of data, drafting of the manuscript, critical revision of the manuscript, final approval of the manuscript.

## Data Sharing

Original data will be shared from the corresponding authors upon reasonable request.

## REFERENCES

1. Fleckenstein, M. et al. Age-related macular degeneration. Nat. Rev. Dis. Prim. 7, 31 (2021).

2. Schmitz-Valckenberg, S. et al. Geographic atrophy: Semantic considerations and literature review. Retina 36, 2250–2264 (2016).

3. Holz, F. G., Strauss, E. C., Schmitz-Valckenberg, S. & Van Lookeren Campagne, M. Geographic atrophy: Clinical features and potential therapeutic approaches. Ophthalmology 121, 1079–1091 (2014).

4. Hughes, S. et al. Prolonged intraocular residence and retinal tissue distribution of a fourth-generation compstatin-based C3 inhibitor in non-human primates. Clin. Immunol. 214, 108391 (2020).

5. Liao, D. S. et al. Complement C3 Inhibitor Pegcetacoplan for Geographic Atrophy Secondary to Age-Related Macular Degeneration: A Randomized Phase 2 Trial. Ophthalmology 127, 186–195 (2020).

6. Apellis Pharmaceuticals. Apellis Announces Top-Line Results from Phase 3 DERBY and OAKS Studies in Geographic Atrophy (GA) and Plans to Submit NDA to FDA in the First Half of 2022.

7. Fleckenstein, M. et al. The Progression of Geographic Atrophy Secondary to Age-Related Macular Degeneration. Ophthalmology 125, 369–390 (2018).

8. Pfau, M. et al. Mesopic and Dark-Adapted Two-Color Fundus-Controlled Perimetry in Geographic Atrophy Secondary to Age-Related Macular Degeneration. Retina (2018). doi:10.1097/IAE.0000000000002337

9. Lindner, M. et al. Combined fundus autofluorescence and near infrared reflectance as prognostic biomarkers for visual acuity in foveal-sparing geographic atrophy. Investig. Ophthalmol. Vis. Sci. 58, BIO61–BIO67 (2017).

10. Chakravarthy, U. et al. Characterizing Disease Burden and Progression of Geographic Atrophy Secondary to Age-Related Macular Degeneration. Ophthalmology 125, 842–849 (2018).

11. Sivaprasad, S. et al. Reliability and Construct Validity of the NEI VFQ-25 in a Subset of Patients With Geographic Atrophy From the Phase 2 Mahalo Study. Am. J. Ophthalmol. 190, 1–8 (2018).

12. Künzel, S. H. et al. Determinants of Quality of Life in Geographic Atrophy Secondary to Age-Related Macular Degeneration. Invest. Ophthalmol. Vis. Sci. 61, 63 (2020).

13. Fleckenstein, M. et al. High-resolution spectral domain-OCT imaging in geographic atrophy associated with age-related macular degeneration. Investig. Ophthalmol. Vis. Sci. 49, 4137–4144 (2008).

14. Li, M. et al. HISTOLOGY OF GEOGRAPHIC ATROPHY SECONDARY TO AGE-RELATED MACULAR DEGENERATION: A Multilayer Approach. Retina 38, 1937–1953 (2018).

15. Spaide, R. F. Outer retinal atrophy after regression of subretinal drusenoid deposits as a newly recognized form of late age-related macular degeneration. Retina 33, 1800–1808 (2013).

16. Chen, L. et al. SUBRETINAL DRUSENOID DEPOSIT IN AGE-RELATED MACULAR DEGENERATION: Histologic Insights Into Initiation, Progression to Atrophy, and Imaging. Retina 40, 618–631 (2020).

17. Steinberg, J. S. et al. Correlation of Partial Outer Retinal Thickness with Scotopic and Mesopic Fundus-Controlled Perimetry in Patients with Reticular Drusen. Am. J. Ophthalmol. 168, 52–61 (2016).

18. Gin, T. J., Wu, Z., Chew, S. K. H., Guymer, R. H. & Luu, C. D. Quantitative analysis of the ellipsoid zone intensity in phenotypic variations of intermediate age-related macular degeneration. Investig. Ophthalmol. Vis. Sci. 58, 2079–2086 (2017).

19. Pfau, M. et al. Mesopic and dark-adapted two-color fundus-controlled perimetry in patients with cuticular, reticular, and soft drusen. Eye 32, 1819–1830 (2018).

20. Pfau, M. et al. Progression of Photoreceptor Degeneration in Geographic Atrophy Secondary to Age-related Macular Degeneration. JAMA Ophthalmol. (2020). doi:10.1001/jamaophthalmol.2020.2914

21. Reiter, G. S. et al. Subretinal Drusenoid Deposits and Photoreceptor Loss Detecting Global and Local Progression of Geographic Atrophy by SD-OCT Imaging. Invest. Ophthalmol. Vis. Sci. 61, 11 (2020).

22. Bird, A. C., Phillips, R. L. & Hageman, G. S. Geographic atrophy: A histopathological assessment. JAMA Ophthalmol. 132, 338–345 (2014).

23. Pfau, M. et al. Determinants of cone- and rod-function in geographic atrophy: AI-based structure-function correlation. Am. J. Ophthalmol. 217, 162–173 (2020).

24. Riedl, S. et al. The effect of pegcetacoplan treatment on photoreceptor maintenance in geographic atrophy monitored by AI-based OCT analysis. Ophthalmol. Retin. (2022). doi:10.1016/j.oret.2022.05.030

25. Thiele, S. et al. Natural history of the relative ellipsoid zone reflectivity in age-related macular degeneration. Ophthalmol. Retin. (2022). doi:10.1016/j.oret.2022.06.001

26. Schmitz-Valckenberg, S. et al. Fundus autofluorescence and fundus perimetry in the junctional zone of geographic atrophy in patients with age-related macular degeneration. Investig. Ophthalmol. Vis. Sci. 45, 4470–4476 (2004).

27. Künzel, S. H. et al. Association of Reading Performance in Geographic Atrophy Secondary to Age-Related Macular Degeneration With Visual Function and Structural Biomarkers. JAMA Ophthalmol. 139, 1191–1199 (2021).

28. Lindner, M. et al. Determinants of Reading Performance in Eyes with Foveal-Sparing Geographic Atrophy. Ophthalmol. Retin. 3, 201–210 (2019).

29. Sunness, J. S. Reading newsprint but not headlines: pitfalls in measuring visual acuity and color vision in patients with bullseye maculopathy and other macular scotomas. Retin. Cases Brief Rep. 2, 83–84 (2008).

30. Sunness, J. S. et al. The Long-term Natural History of Geographic Atrophy from Age-Related Macular Degeneration. Enlargement of Atrophy and Implications for Interventional Clinical Trials. Ophthalmology 114, 271–277 (2007).

31. Pfau, M. et al. Green-Light Autofluorescence Versus Combined Blue-Light Autofluorescence and Near-Infrared Reflectance Imaging in Geographic Atrophy Secondary to Age-Related Macular Degeneration. Invest. Ophthalmol. Vis. Sci. 58, BIO121–BIO130 (2017).

32. Holz, F. G. et al. Efficacy and safety of lampalizumab for geographic atrophy due to age-related macular degeneration: Chroma and spectri phase 3 randomized clinical trials. JAMA Ophthalmol. 136, 666–677 (2018).

33. Schmitz-Valckenberg, S. et al. Semiautomated image processing method for identification and quantification of geographic atrophy in age-related macular degeneration. Investig. Ophthalmol. Vis. Sci. 52, 7640–7646 (2011).

34. Niu, S. et al. Automated retinal layers segmentation in SD-OCT images using dualgradient and spatial correlation smoothness constraint. Comput. Biol. Med. 54, 116–128 (2014).

35. Gorgi Zadeh, S. et al. CNNs enable accurate and fast segmentation of drusen in optical coherence tomography. in Lecture Notes in Computer Science (including subseries Lecture Notes in Artificial Intelligence and Lecture Notes in Bioinformatics) (eds. Cardoso, M. J. et al.) 10553 LNCS, 65–73 (Springer International Publishing, 2017).

36. Fang, L. et al. Automatic segmentation of nine retinal layer boundaries in OCT images of non-exudative AMD patients using deep learning and graph search. Biomed. Opt. Express 8, 2732 (2017).

37. Maloca, P. M. et al. Validation of automated artificial intelligence segmentation of optical coherence tomography images. PLoS One 14, e0220063 (2019).

38. Zweifel, S. A., Spaide, R. F., Curcio, C. A., Malek, G. & Imamura, Y. Reticular Pseudodrusen Are Subretinal Drusenoid Deposits. Ophthalmology 117, 303–12.e1 (2010).

39. Curcio, C. A. & Allen, K. A. Aging of the Human Photoreceptor Mosaic: Evidence for Selective Vulnerability of Rods in Central Retina. Invest. Ophthalmol. Vis. Sci. 34, 3278–3296 (1993).

40. Curcio, C. A., Medeiros, N. E. & Millican, C. L. Photoreceptor loss in age-related macular degeneration. Investig. Ophthalmol. Vis. Sci. 37, 1236–1249 (1996).

41. Hill, A. B. The Environment and Disease: Association or Causation? Proc. R. Soc. Med. 58, 295–300 (1965).

42. Takahashi, A. et al. Photoreceptor Damage and Reduction of Retinal Sensitivity Surrounding Geographic Atrophy in Age-Related Macular Degeneration. Am. J. Ophthalmol. 168, 260–268 (2016).

43. Pfau, M. et al. Light sensitivity within areas of geographic atrophy secondary to agerelated macular degeneration. Investig. Ophthalmol. Vis. Sci. 60, 3992–4001 (2019).

44. Pfau, M. et al. Mesopic And Dark-adapted Two-color Fundus-controlled Perimetry In Geographic Atrophy Secondary To Age-related Macular Degeneration. Retina 40, 169–180 (2020).

45. Wu, Z. et al. Optical coherence tomography-defined changes preceding the development of drusen-associated atrophy in age-related macular degeneration. Ophthalmology 121, 2415–2422 (2014).

46. Wu, Z. et al. Prospective Longitudinal Evaluation of Nascent Geographic Atrophy in Age-Related Macular Degeneration. Ophthalmol. Retin. 4, 568–575 (2020).

47. Guymer, R. H. et al. Incomplete Retinal Pigment Epithelial and Outer Retinal Atrophy in Age-Related Macular Degeneration: Classification of Atrophy Meeting Report 4. Ophthalmology 127, 394–409 (2020).

48. Wu, Z. et al. OCT Signs of Early Atrophy in Age-Related Macular Degeneration: Interreader Agreement: Classification of Atrophy Meetings Report 6. Ophthalmol. Retin. 6, 4–14 (2022).

49. Nittala, M. G. et al. Association of Pegcetacoplan With Progression of Incomplete Retinal Pigment Epithelium and Outer Retinal Atrophy in Age-Related Macular Degeneration: A Post Hoc Analysis of the FILLY Randomized Clinical Trial. JAMA Ophthalmol. (2022). doi:10.1001/jamaophthalmol.2021.6067

50. Nittala, M. G. et al. Association of Pegcetacoplan With Progression of Incomplete Retinal Pigment Epithelium and Outer Retinal Atrophy in Age-Related Macular Degeneration: A Post Hoc Analysis of the FILLY Randomized Clinical Trial. JAMA Ophthalmol. (2022). doi:10.1001/jamaophthalmol.2021.6067

51. Sadigh, S. et al. Abnormal thickening as well as thinning of the photoreceptor layer in intermediate age-related macular degeneration. Investig. Ophthalmol. Vis. Sci. 54, 1603–1612 (2013).

52. Lu, C. D. et al. Photoreceptor layer thickness changes during dark adaptation observed with ultrahigh-resolution optical coherence tomography. Investig. Ophthalmol. Vis. Sci. 58, 4632–4643 (2017).

53. Jaffe, G. J. et al. C5 Inhibitor Avacincaptad Pegol for Geographic Atrophy Due to Age-Related Macular Degeneration: A Randomized Pivotal Phase 2/3 Trial. Ophthalmology 128, 576–586 (2021).

54. Jaffe, G. J. et al. Development of IONIS-FB-LRx to treat geographic atrophy associated with AMD. Invest. Ophthalmol. Vis. Sci. 61, 4305 (2020).

55. Khanani, A. M. et al. Phase 1 Study of the Anti-HtrA1 Antibody-binding Fragment FHTR2163 in Geographic Atrophy Secondary to Age-related Macular Degeneration. Am. J. Ophthalmol. 232, 49–57 (2021).

56. R Core Team. R: A Language and Environment for Statistical Computing. (2021).

57. Wickham, H., François, R., Henry, L. & Müller, K. dplyr: A Grammar of Data Manipulation. (2021).

58. Valero-Mora, P. M. ggplot2: Elegant Graphics for Data Analysis. Journal of Statistical Software 35, (Springer, 2010).

59. Bates, D., Mächler, M., Bolker, B. & Walker, S. Fitting Linear Mixed-Effects Models Using lme4. J. Stat. Software; Vol 1, Issue 1 (2015).

60. Lenth, R. V. emmeans: Estimated Marginal Means, aka Least-Squares Means. (2021).

61. Martin Bland, J. & Altman, D. G. Statistical Methods for Assessing Agreement Between Two Methods of Clinical Measurement. Lancet 327, 307–310 (1986).

